# Identifying probable dementia in undiagnosed Black and White Americans using machine learning in Veterans Health Administration electronic health records

**DOI:** 10.1101/2023.02.08.23285540

**Authors:** Yijun Shao, Kaitlin Todd, Andrew Shutes-David, Steven P. Millard, Karl Brown, Amy Thomas, Kathryn Chen, Katherine Wilson, Qing T. Zeng, Debby W. Tsuang

## Abstract

The application of machine learning (ML) tools in electronic health records (EHRs) can help reduce the underdiagnosis of dementia, but models that are not designed to reflect minority population may perpetuate that underdiagnosis. To address the underdiagnosis of dementia in both Black Americans (BAs) and white Americans (WAs), we sought to develop and validate ML models that assign race-specific risk scores. These scores were used to identify undiagnosed dementia in BA and WA Veterans in EHRs. More specifically, risk scores were generated separately for BAs (n=10K) and WAs (n=10K) in training samples of cases and controls by performing ML, equivalence mapping, topic modeling, and a support vector-machine (SVM) in structured and unstructured EHR data. Scores were validated via blinded manual chart reviews (n=1.2K) of controls from a separate sample (n=20K). AUCs and negative and positive predictive values (NPVs and PPVs) were calculated to evaluate the models. There was a strong positive relationship between SVM-generated risk scores and undiagnosed dementia. BAs were more likely than WAs to have undiagnosed dementia per chart review, both overall (15.3% vs 9.5%) and among Veterans with >90^th^ percentile cutoff scores (25.6% vs 15.3%). With chart reviews as the reference standard and varied cutoff scores, the BA model performed slightly better than the WA model (AUC=0.86 with NPV=0.98 and PPV=0.26 at >90^th^ percentile cutoff vs AUC=0.77 with NPV=0.98 and PPV=0.15 at >90^th^). The AUCs, NPVs, and PPVs suggest that race-specific ML models can assist in the identification of undiagnosed dementia, particularly in BAs. Future studies should investigate implementing EHR-based risk scores in clinics that serve both BA and WA Veterans.

## 1 Introduction

Alzheimer’s disease (AD) and related dementias (ADRD) are fatal neurodegenerative disorders that account for half of admissions to long-term care facilities (Rice et al., 2001), yet nearly half of those affected by ADRD have not been formally diagnosed (Barnes et al., 2020, Amjad et al., 2018). This crisis of underdiagnosis exacerbates existing disparities in health care, as dementia underdiagnosis may disproportionately affect Black Americans (BAs) (Gianattasio et al., 2019). In a large 2019 study of Medicare claims, older BAs with dementia were about two times less likely to be correctly diagnosed with dementia than older White Americans (WAs) with dementia (Gianattasio et al., 2019), and in one of the small handful of studies that examine racial disparity in dementia care within VHA (Sleath et al., 2005, Kalkonde et al., 2009), significantly fewer BA Veterans with suspected dementia underwent neuropsychological testing for the diagnosis of dementia than WA Veterans with suspected dementia (Kalkonde et al., 2009). The underdiagnosis of dementia translates into missed opportunities to treat patients (Cummings et al., 2021), improve quality of life (e.g., through medication management and referrals) (Callahan et al., 1995, Fitten et al., 1995), reduce patient and family burden (Sayegh and Knight, 2013, Hinton et al., 2004), and reduce hospitalization, institutionalization, and health care costs (Rasmussen and Langerman, 2019, Black et al., 2018).

We seek to use natural language processing (NLP) and machine learning (ML) tools to address the magnitude of dementia diagnostic disparity in the Veterans Health Administration (VHA) Corporate Data Warehouse (CDW), which is an ideal setting for this work, as it contains comprehensive structured and unstructured data on ∼0.4 million BA Veterans who are age 65+ and receive care as part of the largest integrated health care system in the nation. ML methods have previously been applied to EHRs (Nadkarni et al., 2011, Gottesman et al., 2013), but we have developed one of the *first* ML models to increase the sensitivity of dementia identification by using *both* structured EHR data (e.g., demographics, diagnoses [ICD codes], procedures [CPTS codes], medications, and clinical note types) and unstructured EHR data (e.g., words in clinical notes) (Shao et al., 2019). In our previous work, we applied topic modeling and logistic regression to develop risk scores for dementia based on the EHRs of older Veterans with (n=1,861, mean age 79.8) and without (n=9,305, mean age 79.5) ICD-9 dementia codes (Shao et al., 2019). Here, we extend this work by building separate predictive models for detecting undiagnosed dementia in BAs and WAs using a larger sample of all VA patients who are 65+ years old with and without ICD 9/10 diagnosed dementia. We validate these models by performing chart reviews blinded to dementia risk scores in a new set of patients who lack ICD-9/10 dementia diagnoses and who were not used to build the models; we then compare the chart review diagnoses to the diagnoses based on the model-generated risk scores.

## 2 Materials and Methods

### 2.1 Study population

After receiving IRB approval, we created a cohort of cases (i.e., Veterans with an ICD-9/10 dementia code) and controls (Veterans without any ICD-9/10 dementia codes) from the CDW by selecting patients who turned age 65 between 1999 and 2018, lacked a dementia diagnosis at age 65, were previously evaluated at a VA clinic, and were identified as BA or WA in their EHRs (top row, Figures 1a and 1b). The selected Veterans were followed until 9/12/2018, until diagnosis (cases), or until censoring due to absence of records (controls).

**Figure 1a.**
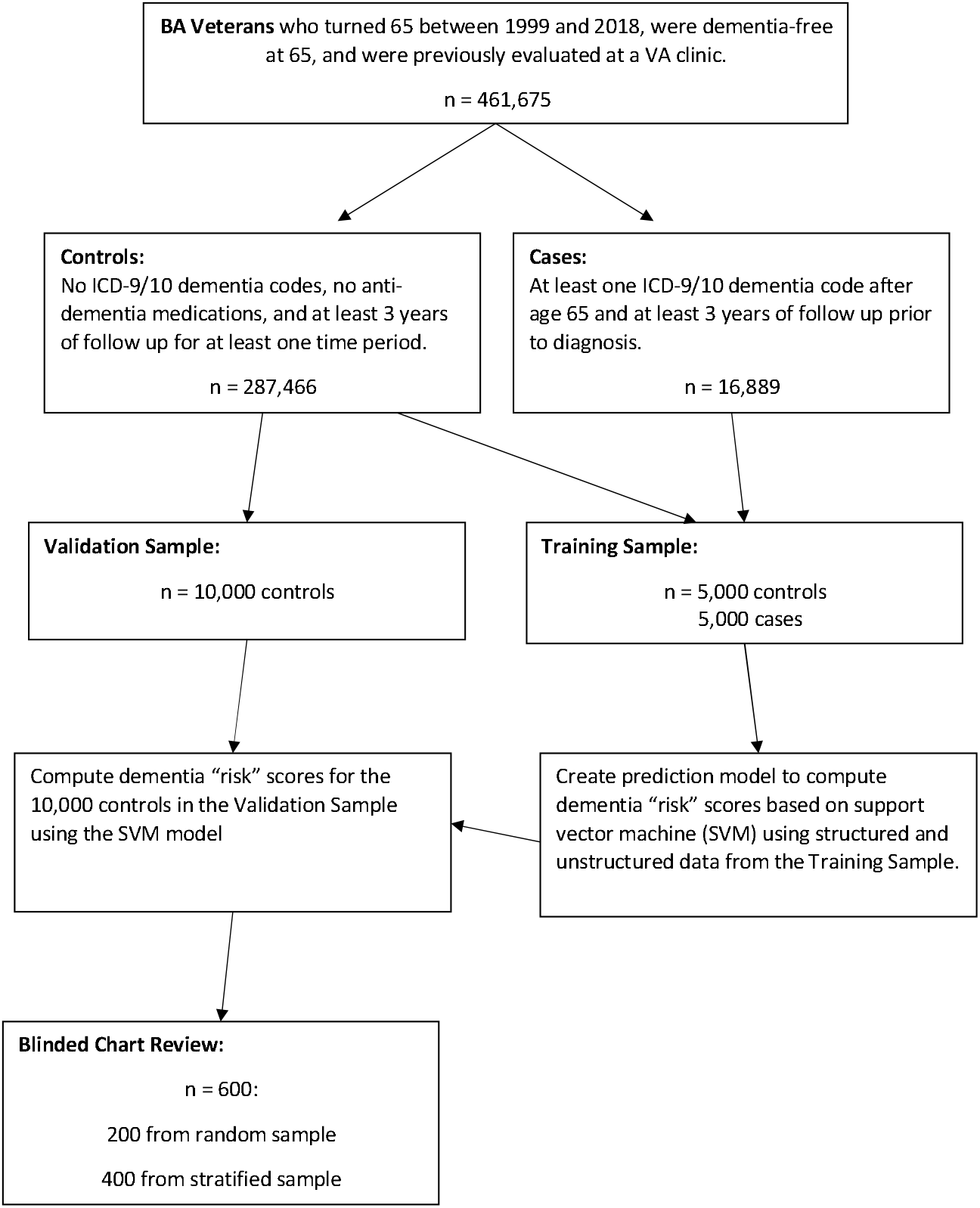
Study flow diagram for Black American (BA) Veterans. This figure shows the number of BA Veterans available within the Veterans Health Administration (VHA) Corporate Data Warehouse (CDW) for the time period under study who met inclusion/exclusion criteria, as well as the number of Veterans used for model building and validation. Veterans in the Training Sample and Validation Sample were chosen with simple random sampling. Veterans who underwent chart review (blinded to score) were chosen from the 10,000 in the Validation Sample by simple random sampling (n = 200) and stratified random sampling (n = 400), where the strata were based on the scores.

**Figure 1b.**
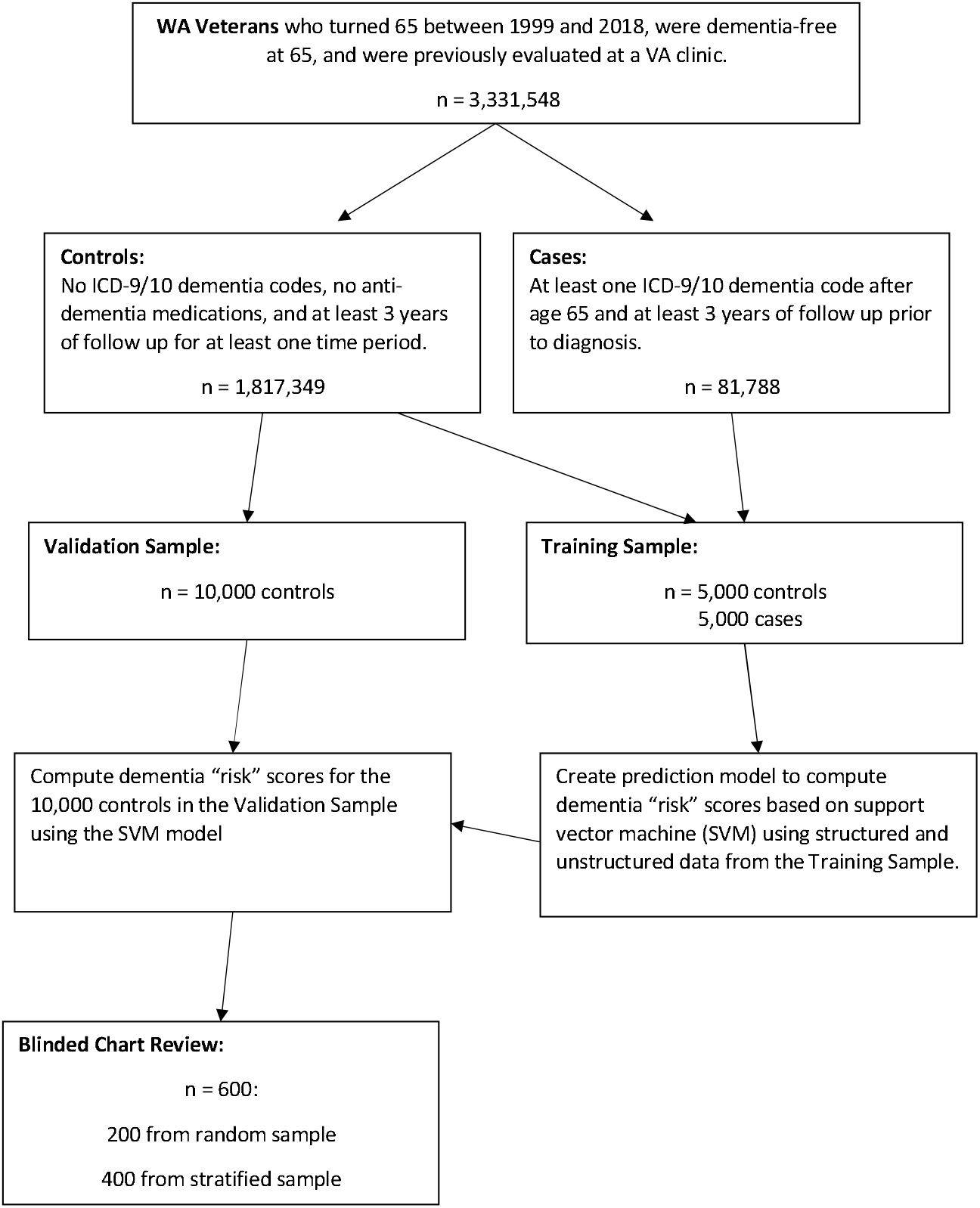
Study flow diagram for White American (WA) Veterans. This figure shows the number of WA Veterans available within the Veterans Health Administration (VHA) Corporate Data Warehouse (CDW) for the time period under study who met inclusion/exclusion criteria, as well as the number of Veterans used for model building and validation. Veterans in the Training Sample and Validation Sample were chosen with simple random sampling. Veterans who underwent chart review (blinded to score) were chosen from the 10,000 in the Validation Sample by simple random sampling (n = 200) and stratified random sampling (n = 400), where the strata were based on the scores.

To meet inclusion criteria, cases had to have received at least one ICD-9 or ICD-10 diagnosis of dementia, with the first diagnosis occurring after age 65, and had to have at least 3 years of continuous follow-up (i.e., 2+ documented clinical visits and associated notes during each year) immediately prior to first diagnosis. That is, the one-year-long period in which first diagnosis occurred had to have at least 3 visits (i.e., a diagnosis visit plus 2 previous visits), whereas the other 2 one-year-long periods had to have at least 2 visits. Conversely, controls could not have had any ICD-9/10 dementia codes; could not have filled donepezil, galantamine, rivastigmine, or memantine prescriptions; and needed 3+ years of continuous follow-up (i.e., 2+ documented clinical visits and associated notes during each year) after reaching age 62. We created separate BA and WA cohorts of cases and controls to satisfy these criteria (second row, Figures 1a and 1b).

All clinical data were collected for a 3-year period that either immediately preceded but did not include the first ICD-9/10 diagnosis of dementia (for cases) or a random visit date that was selected as an index date (for controls). This 3-year period was established to provide adequate structured and unstructured data.

The sampling and modeling of the Training and Validation Samples was performed separately for BAs and WAs. We created model Training Samples by randomly sampling 5,000 cases and 5,000 controls in each race (total n=20,000). For each control, we randomly chose the index visit among all visits that satisfied the 3-year lookback criterion. We used the Training Samples to build models that produced dementia risk scores. We then created model Validation Samples by randomly sampling 10,000 controls in each race who were not part of the Training Samples (total n=20,000) and used the models to generate scores for these samples. Finally, we sampled 600 Veterans from the Validation Samples for each race to undergo blinded chart reviews (total n=1,200). Veterans were selected for chart review by simple random sampling (n=200) and stratified random sampling (n=400) based on percentiles of the full Validation Sample risk scores, such that 100 Veterans from the >75^th^– 90^th^ percentiles were included, and 30 Veterans in each of the 10 remaining upper percentile ranges (i.e., 30 each from the >90^th^– 91^st^, >91^st^–92^nd^, etc.) were included.

### 2.2 Variable creation

#### 2.2.1 Structured data

For each Veteran, we aggregated the structured data over the 3-year analysis period, recording the presence/absence of each type of structured data during the 3-year period. Each type of structured data was treated as a candidate binary variable for our model that would produce dementia risk scores, with 0 indicating an absence of the codes/medications/note type and 1 indicating their presence.

To account for a transition from ICD-9 to ICD-10, we performed equivalence mapping, visualizing the CDC/CMS general equivalence mappings (GEM) as a large bipartite graph that consisted of two disjointed sets of vertices representing all the ICD-9 and ICD-10 codes, respectively, and a number of edges connecting ICD-9 vertices to ICD-10 vertices representing the possible conversions from ICD-9 codes to ICD-10 codes. These mappings allowed us to decompose the GEM, viewed as a large bipartite graph, into a number of smaller disjoint bipartite subgraphs that could not be decomposed into smaller disjoint subgraphs without breaking edges. Then, for each of these minimal equivalence mappings, a new code was defined to represent the group of ICD-9 codes before the transition date and the group of ICD-10 codes after the transition. Variables corresponding to the new codes were defined in the same way as other codes (e.g., CPT codes).

#### 2.2.2 Unstructured data

Unstructured data were handled using the two-step topic modeling approach previously described in Shao et al. (Shao et al., 2016, Shao et al., 2019). This unsupervised ML method identifies shared topics from a large text corpus. Each topic is defined as a binary variable indicating the presence/absence of that topic, and the proportion of topics within any particular document is calculated. Here, we use the proportion of dementia-related topics observed in excess in cases versus controls to identify dementia-related signs.

More specifically, raw topics were identified in clinical notes by running a latent Dirichlet allocation (LDA) algorithm within the Machine Learning for Language Toolkit Java package (Shao et al., 2016, Shao et al., 2019), which includes topic learning and inference functions. The learning function is a time-consuming algorithm that learns the topics from a set of text documents and generates a topic model, whereas the inference function runs much faster and can apply the learned topic model to a new set of text documents and then infer the topic distributions in those documents. For our topic learning subset, we randomly sampled one note per day for each subject from the ∼5 million notes collected during the 3-year study period, yielding a sample corpus of 1.8 million notes. We randomly selected 1 million notes from this sample corpus, which allowed for a reduced running time for topic learning while ensuring that main topics were preserved. We then ran LDA topic learning 3 times on the 1 million sampled notes, setting 1,000 as the total number of topics, and applied the 3 resulting models to all of the 5 million notes, using the topic inference function to infer the topic proportions in each note. Based on the inferred topic proportions, we calculated the number of words that were associated with each topic in each note by multiplying the topic proportion by the total number of words in the note. Because the “number of words” associated with a topic was not always a whole number, we call it the pseudo word count (PWC).

We then applied the stable topic extraction method (Shao et al., 2016, Shao et al., 2019), which yielded 852 stable topics. For each stable topic, there were 3 topics—one from each run— that were very similar to each other, and the stable topic was the “average” of the 3 similar topics. Likewise, the PWC for the stable topic in each note was defined to be the median value of the 3 PWCs corresponding to the 3 topics. By design, topic proportions are always positive numbers, so the PWCs are positive as well. However, because not all of the topics are present in every note, we set a nonzero threshold on the PWCs to indicate whether a topic was present in a note. Empirically, we set the threshold at 2.0, which roughly means that a topic is present in a note only when the PWC≥2.0. To allow various degrees of topic presence, we defined topic presence to be a function of PWC as follows: (1) presence=0 if PWC<2.0, (2) presence=PWC/10.0 if 2.0≤PWC≤10.0, and (3) presence=1.0 if PWC>10.0. For the ML model, stable topics were used as variables/features, and the maximum presence value over all the notes of each Veteran was defined as the Veteran’s topic presence value.

### 2.3 Variable selection

Separately for BAs and WAs, we selected variables from the structured data that corresponded to the codes/medications/note types that were present in 10+ Veterans in the Training Sample. All of the stable topic variables and two demographic variables (age and sex) were selected. The age variable was normalized so that the value 0 corresponded to 65 years old (minimum age) whereas the value 1 corresponded to 85 years old (maximum age). All other variables were either binary (i.e., values 0 and 1) or continuous (i.e., values between 0 and 1).

### 2.4 Support vector machine (SVM) model

Separately for BA and WA Veterans in the Training Sample, we constructed SVM models that used the selected predictor variables to generate dementia “risk” scores. To construct the SVM models, we used the linear SVM model (LinearSVC algorithm) in Python package *scikit-learn* (Pedregosa et al., 2011). The SVM models had only one important hyperparameter: “C,” the cost parameter, which sets the trade-off between misclassification and the simplicity of the decision surface. To determine the best value for C, we performed five-fold cross-validation on the training dataset with various values for C and then selected the value corresponding to the highest predictive area under the receiver operating characteristic (ROC) curve (AUC) in the five-fold cross-validation. The selected C value was used to train the final SVM model on the entire training dataset. The linear SVM model output scores represent the distance to the separation hyperplane in the high-dimensional feature space. The scores have no theoretical limits, and higher scores mean indicate a higher likelihood of having dementia.

### 2.5 Validation of the SVM model

We separately generated scores for BA and WA controls in the Validation Sample and then, in a subset of these Veterans, we performed chart reviews in which reviewers were blinded to score. Chart reviews were conducted by experienced cognitive disorder experts (i.e., 2 trained in geriatric psychiatry [DT and KC] and 1 in geriatric medicine [AT]) who achieved interrater reliability on dually reviewed charts (Cohen’s Kappa value of 0.74 [se = 0.25, 95% CI = 0.25 - 1; p = 0.0016]). The reviewers retroactively applied the DSM-V criteria for major neurocognitive disorder (Sachdev et al., 2014) by evaluating memory, apraxia, aphasia, agnosia, executive functioning, and functional domains of ADL and iAD (Katz, 1983) in abstracted notes. Reviewers avoided attributing cognitive or functional deficits due to physical limitations or acute or chronic medical conditions to dementia. When reviewers were uncertain about a Veteran’s dementia status, that Veteran was labeled *uncertain* and then one of the other reviewers adjudicated dementia status independent of the initial reviewer. Dementia status was coded by reviewers as “None,” “Possible,” or “Probable”; a probable or possible dementia code thus indicated that a Veteran had dementia symptoms that had either not been worked up nor previously assigned a dementia diagnosis. Using chart review as the reference standard, we assessed the prevalence of undiagnosed dementia and assessed the sensitivity, specificity, positive predictive value (PPV), negative predictive value (NPV), and AUC by varying the cutoff score for determining when to declare “possible or probable undiagnosed dementia.” Estimates were computed using inverse probability weighting to account for stratified sampling (Alonzo and Pepe, 2005), and confidence intervals were computed using bootstrapping. Demographics, estimates, and confidence intervals were computed using R (R Core Team, 2020). We created scatter plots of dementia risks for 3 groups (probable, possible and none) as well as 2 groups (probable/possible combined and none).

## 3 Results

### 3.1 Demographics

Among the Veterans who met inclusion/exclusion criteria (see Figures 1a and 1b), the prevalence of dementia was 5.5% for BAs and 4.3% for WAs. Veterans ranged in age from 65 to 84 (see demographics in Table 1). In the Training Sample, cases were older compared to controls (mean [SD]=72.4 [4.8] vs. 69.1 [3.7]), and both cases and controls were overwhelmingly male (97.7 % and 97.2%). BA Veterans were slightly younger than WA Veterans (72.1 [4.8] vs. 72.8 [4.8] for cases; 68.6 [3.5] vs. 69.5 [3.8] for controls). The demographics for controls in the Validation and Training Sample were similar.

**Table 1a.** Demographics of the Training Sample by Race (BA: Black American; WA: White American).

| Characteristic | Case (n = 10K) |  |  | Control (n = 10K) |  |  |
| --- | --- | --- | --- | --- | --- | --- |
|  | BA (n = 5K) | WA (n = 5K) | Combined (n = 10K) | BA (n = 5K) | WA (n = 5K) | Combined (n = 10K) |
| Age, mean (SD) | 72.1 (4.8) | 72.8 (4.8) | 72.4 (4.8) | 68.6 (3.5) | 69.5 (3.8) | 69.1 (3.7) |
| Age category, (%) |  |  |  |  |  |  |
| 65–69 | 35.7 | 29.8 | 32.8 | 70.5 | 60.0 | 65.2 |
| 70–74 | 34.1 | 34.4 | 34.3 | 22.1 | 28.4 | 25.3 |
| 75–79 | 20.9 | 24.2 | 22.6 | 5.8 | 9.2 | 7.5 |
| 80–84 | 9.4 | 11.5 | 10.4 | 1.6 | 2.5 | 2.0 |
| Gender, % male | 97.9 | 97.5 | 97.7 | 96.8 | 97.6 | 97.2 |

**Table 1b.**
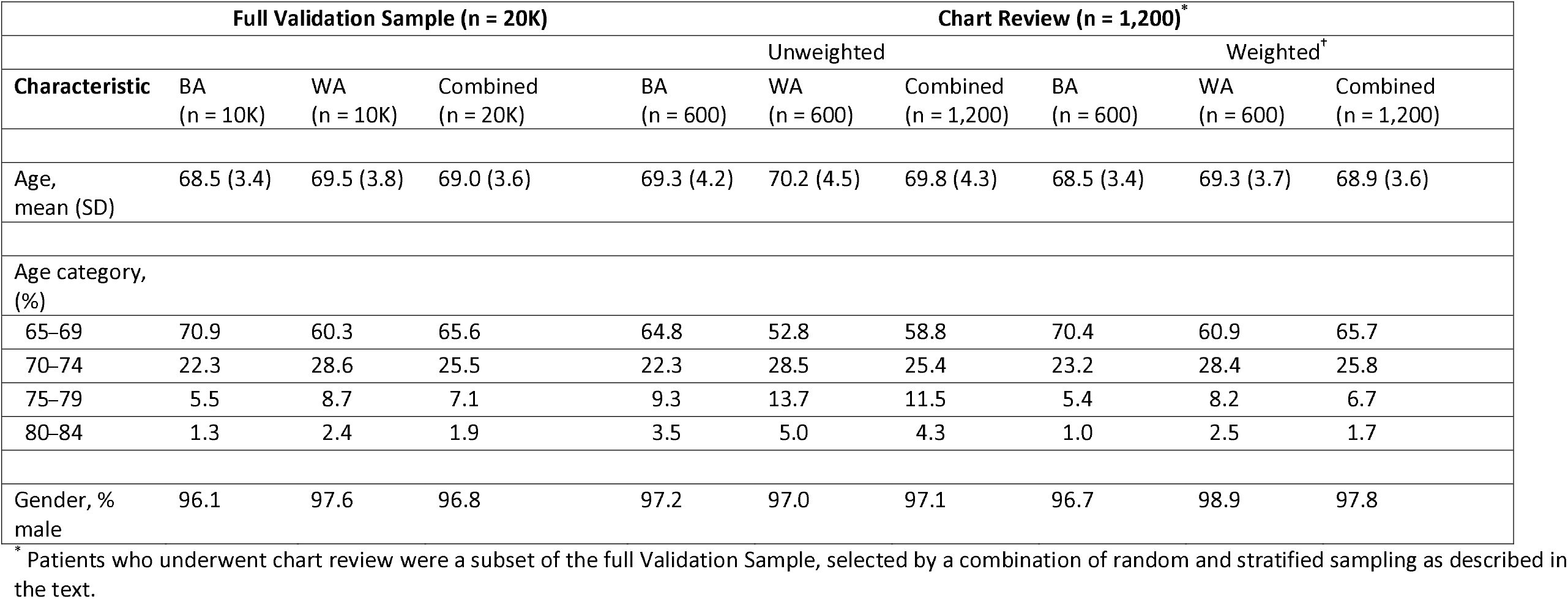
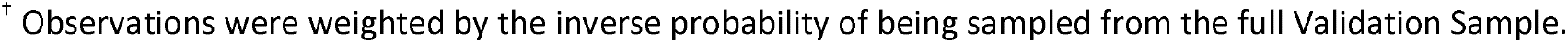
Demographics of the Validation Sample by Race (BA: Black American; WA: White American).

| Characteristic | Full Validation Sample (n = 20K) |  |  | Chart Review (n = 1,200)* |  |  |  |  |  |
| --- | --- | --- | --- | --- | --- | --- | --- | --- | --- |
|  | BA<br>(n = 10K) | WA<br>(n = 10K) | Combined<br>(n = 20K) | Unweighted |  |  | Weighted† |  |  |
|  |  |  |  | BA<br>(n = 600) | WA<br>(n = 600) | Combined<br>(n = 1,200) | BA<br>(n = 600) | WA<br>(n = 600) | Combined<br>(n = 1,200) |
| Age, mean (SD) | 68.5 (3.4) | 69.5 (3.8) | 69.0 (3.6) | 69.3 (4.2) | 70.2 (4.5) | 69.8 (4.3) | 68.5 (3.4) | 69.3 (3.7) | 68.9 (3.6) |
| Age category, (%) |  |  |  |  |  |  |  |  |  |
| 65–69 | 70.9 | 60.3 | 65.6 | 64.8 | 52.8 | 58.8 | 70.4 | 60.9 | 65.7 |
| 70–74 | 22.3 | 28.6 | 25.5 | 22.3 | 28.5 | 25.4 | 23.2 | 28.4 | 25.8 |
| 75–79 | 5.5 | 8.7 | 7.1 | 9.3 | 13.7 | 11.5 | 5.4 | 8.2 | 6.7 |
| 80–84 | 1.3 | 2.4 | 1.9 | 3.5 | 5.0 | 4.3 | 1.0 | 2.5 | 1.7 |
| Gender, % male | 96.1 | 97.6 | 96.8 | 97.2 | 97.0 | 97.1 | 96.7 | 98.9 | 97.8 |

\* Patients who underwent chart review were a subset of the full Validation Sample, selected by a combination of random and stratified sampling as described in the text.
<sup>†</sup> Observations were weighted by the inverse probability of being sampled from the full Validation Sample.

### 3.2 Variable selection for the SVM model

For the model trained on BA Veterans, a total of 8221 features were selected, including 2 demographics, 854 topics, 2229 nondementia ICD code groups, 2561 CPT codes, 686 medications, and 1889 note types. For the model trained on WA Veterans, a total of 7716 features were selected, including 2 demographics, 854 topics, 2141 nondementia ICD code groups, 2330 CPT codes, 655 medications, and 1734 note types.

The most significant topic features are shown in Supplemental Table 1. Note that the terms in a topic can occur in any order or combination, and the presence of a topic in a document does not require that all the terms in a topic be present. Topics that were observed more frequently in cases than in controls were considered dementia related.

### 3.3 Distribution of scores

In the Training Sample, cases had higher scores than controls (mean [SD]=0.56 [0.54] vs. -0.50 [0.36] for BAs and 0.54 [0.55] vs. -0.47 [0.34] for WAs; Figure 2, Supplemental Figure 1). In the Validation Sample, among Veterans with undiagnosed dementia who underwent chart review, those diagnosed by reviewers with possible/probable dementia had higher scores compared to those diagnosed with no dementia (0.45 [0.38] vs. -0.02 [0.51] for BAs, and 0.38 [0.41] vs. -0.02 [0.47] for WAs; Figure 3). For our chart review subsample of the Validation Sample, we oversampled Veterans with higher scores (i.e., Veterans with chart reviews had higher scores compared to all Validation Veterans: 0.05 [0.52] vs. -0.45 [0.41] for BA Veterans, and 0.02 [0.48] vs. -0.44 [0.38] for WA Veterans; Supplemental Figure 2), and therefore, we adjusted scores using inverse probability weighting to account for stratified sampling.

**Figure 2.**
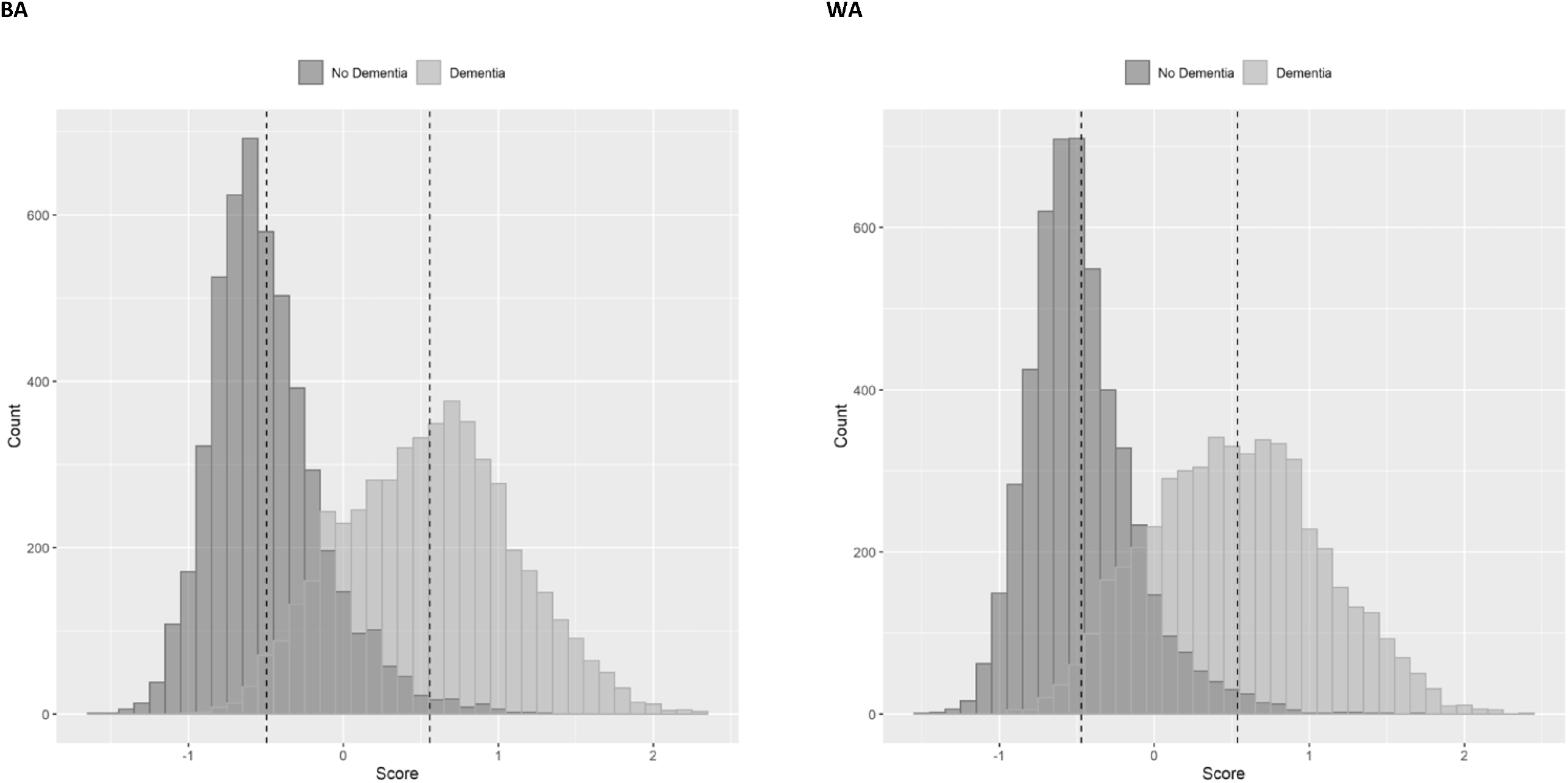
Distribution of scores by dementia status and race (BA: Black American; WA: White American) for Veterans in the Training Sample (n = 5,000 in each dementia status group for each race). Dashed lines represent the means of the distribution.

**Figure 3a:**
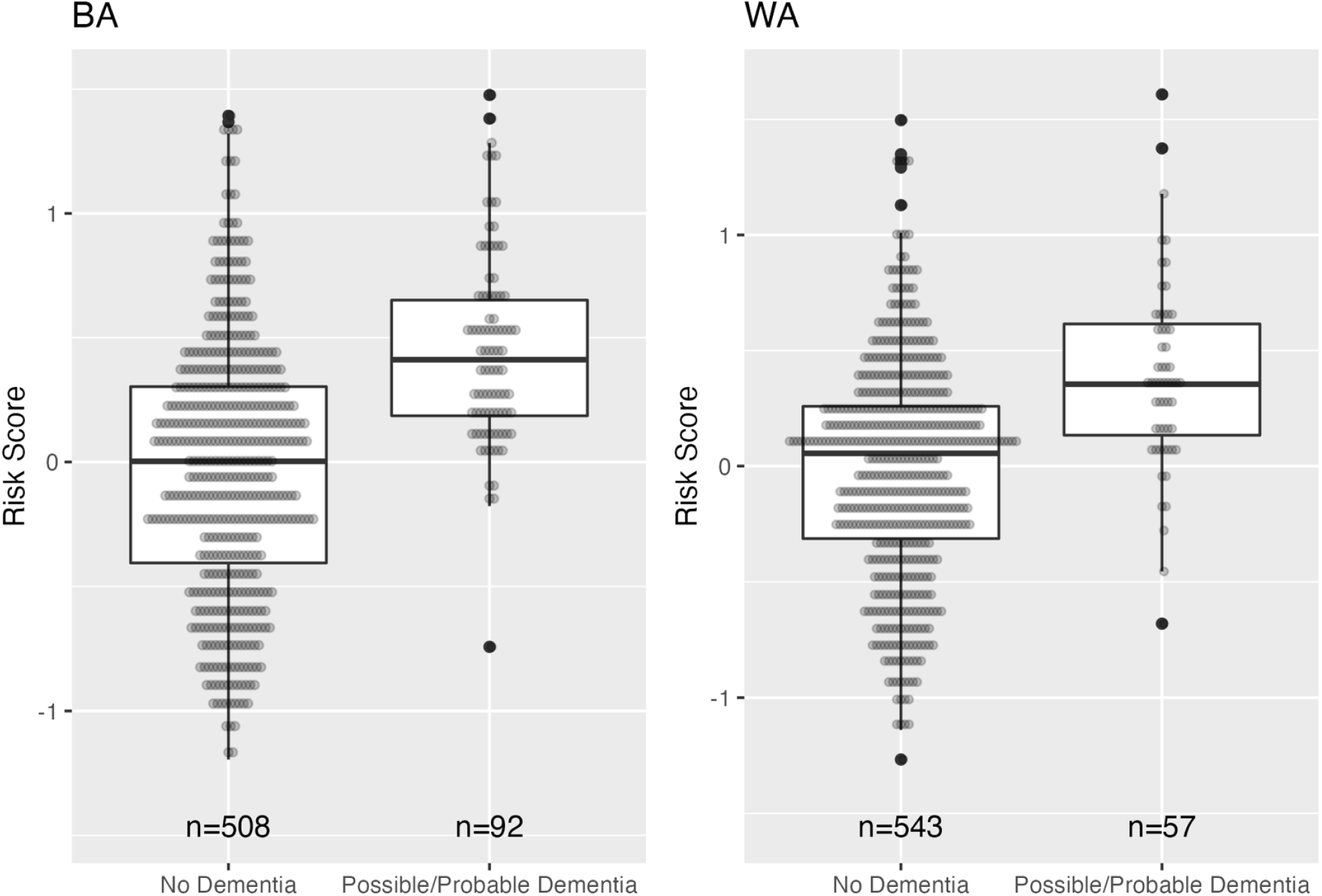
Distribution of risk scores by dementia status and race (BA: Black American, WA: White American) for Veterans in in the Validation Sample who underwent chart review (n = 600 for each race).

**Figure 3b.**
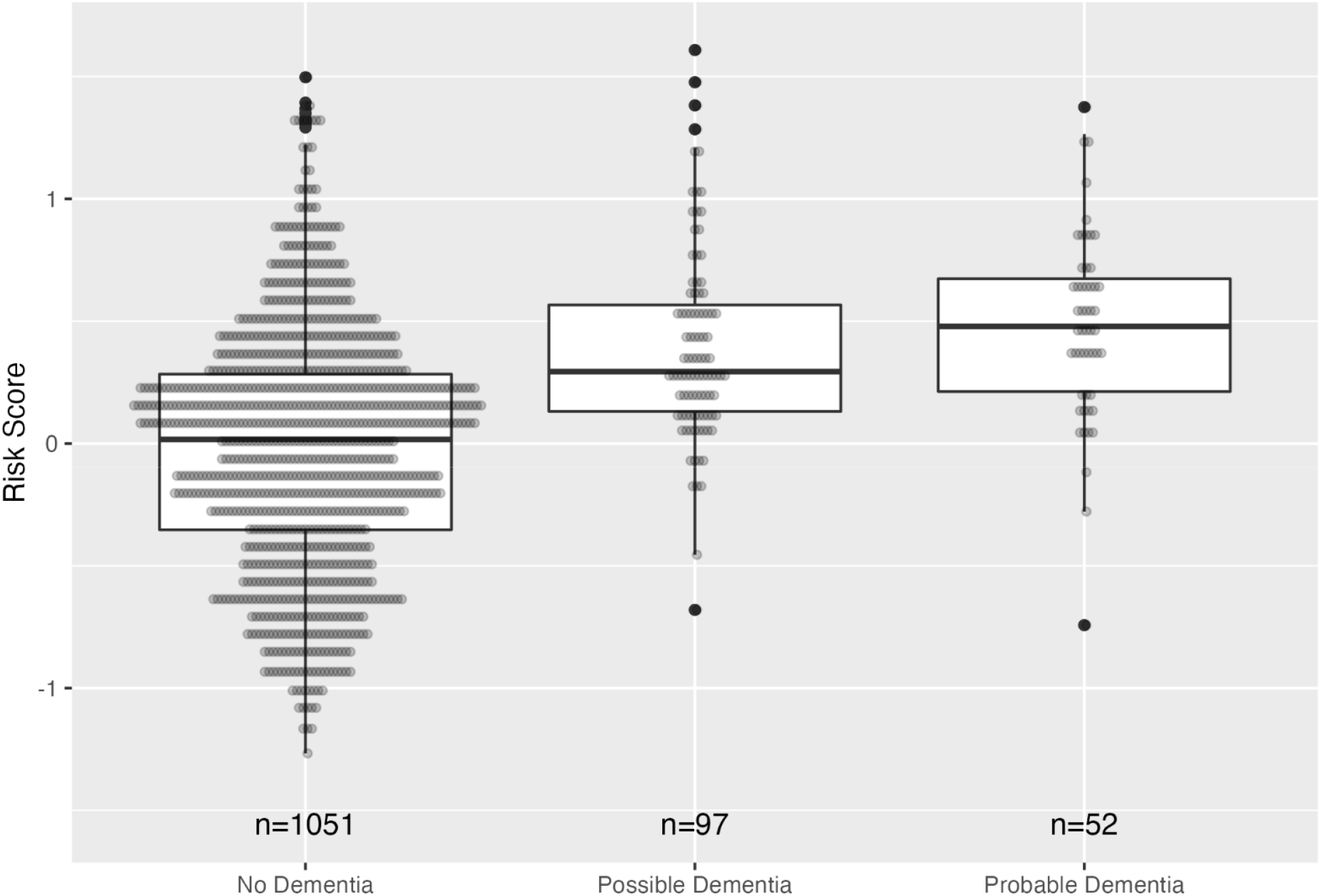
Distribution of risk scores by dementia status for both Black American and White American Veterans in the Validation Sample who underwent chart ew (n = 600 for each race).

### 3.4 Prevalence of undiagnosed dementia and screening test characteristics

Of the 1,200 Veterans who underwent chart review, 15.3% (n=92) of BAs and 9.5% (n=57) of WAs were identified with possible/probable dementia. After adjusting for stratified sampling that intentionally oversampled Veterans with higher scores, the estimated prevalence of undiagnosed dementia in the full Validation Sample was 4.1% [3.2, 6.2] for BA Veterans and 3.6% [2.3, 6.3] for WA Veterans. There was a strong positive relationship between risk scores and the prevalence of undiagnosed dementia (Figure 4), and as anticipated, for Veterans with scores below the 90^th^ percentile, the percentages of undiagnosed dementia were low: 3.9% (95% CI [2.1, 7.0]) and 2.9% (95% CI [1.3, 5.8]) for BA and WA Veterans, respectively. Among Veterans with scores above the 90^th^ percentile, we found that a higher percentage of BA Veterans had undiagnosed dementia than WA Veterans: 25.6% (95% CI [20.9, 30.8]) vs. 15.3% (95% CI [11.6, 19.8]).

**Figure 4.**
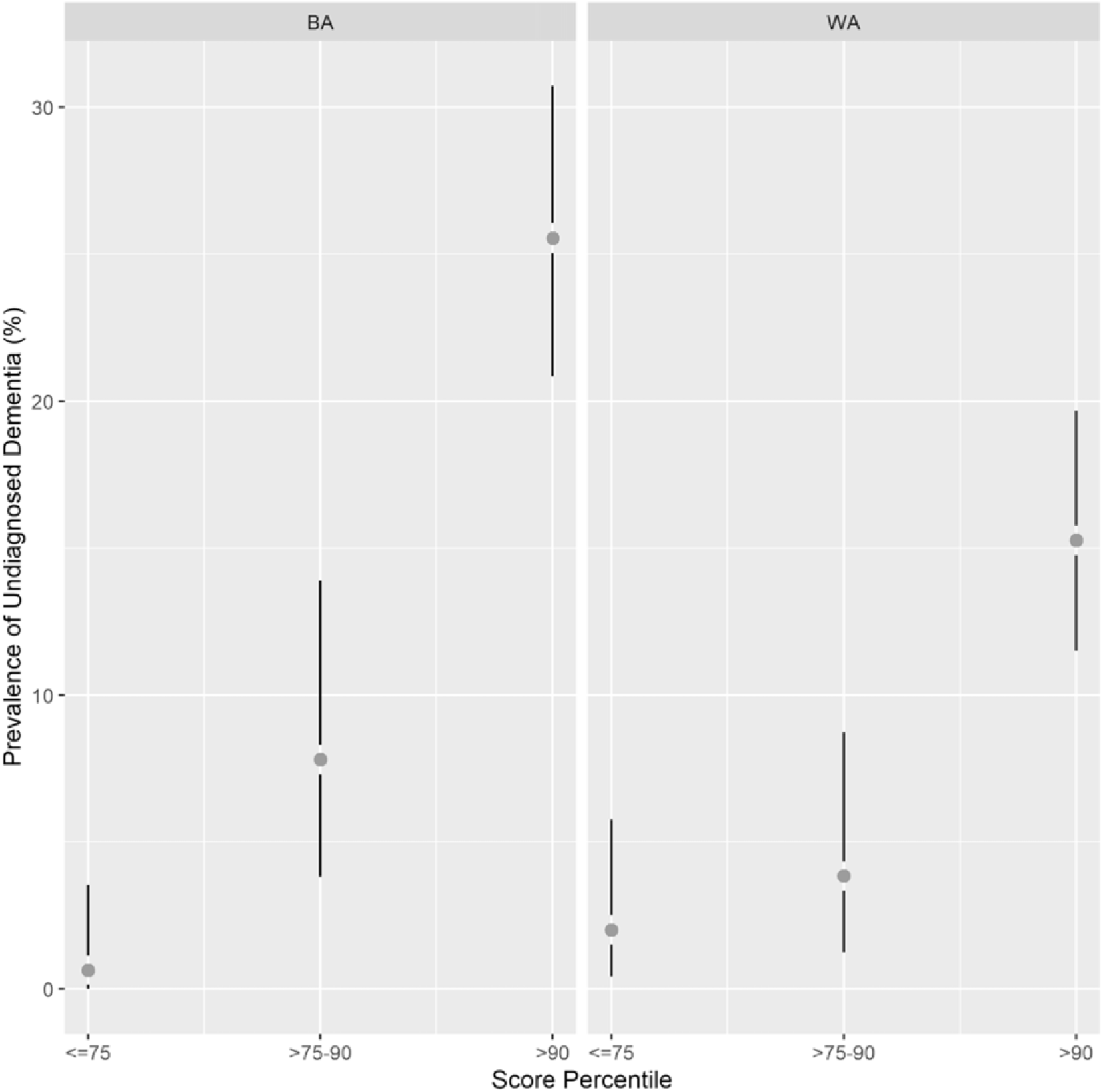
Prevalence of undiagnosed dementia by score percentile stratum and race (BA: Black American; WA: White American) for Veterans who underwent chart review (n = 600 for each race). For each race, score percentiles are based on using the scores from all 10,000 Veterans in the Validation Sample.

Supplemental Figure 3 shows observed values for sensitivity, specificity, PPV, and NPV of the screening tests that use chart review as the reference standard and vary cutoff score, and Supplemental Table 2 lists values for various score cutoffs. As shown in Supplemental Figure 4, the AUC was moderately high for both BA Veterans (0.86 [0.59, 0.95]) and WA Veterans (0.77 [0.59, 0.90]). For score cutoffs above the 50^th^ percentile in the Validation Sample, sensitivity was moderate and specificity was very high for both BA and WA Veterans (e.g., using the 90^th^ percentile as the cutoff, sensitivity and specificity were 0.61 [0.40, 0.76] and 0.92 [0.91, 0.92], respectively, for BA Veterans and 0.43 [0.24, 0.67] and 0.91 [0.91, 0.92], respectively, for WA Veterans). Because of the low prevalence of undiagnosed dementia in the full Validation Samples, as well as the low sensitivity and high specificity of the screening tests, it was unsurprising that PPV was low and NPV was high (Tenny and Hoffman, 2022); using the 90^th^ percentile as the cutoff, PPV was only 0.26 [0.21, 0.30] and 0.15 [0.12, 0.20] for BA and WA Veterans, respectively. In contrast, NPVs remained quite high regardless of the score cutoff.

## 4 Discussion

### 4.1 Significance

We have successfully developed and validated a ML model to identify probable dementia cases in BA and WA Veterans without ICD diagnoses. The dementia risk scores generated by the SVM models were positively correlated with the diagnosis of dementia and achieved a high AUC (0.86 [0.59, 0.95]) for BA Veterans and a satisfactory AUC for WA Veterans (0.77 [0.59, 0.90]). Given that BAs are about twice as likely to develop dementia as WAs (Tang et al., 2001, Langa et al., 2017), the good performance of the SVM in this population is particularly important.

### 4.2 Context

Our preliminary data suggest that BA Veterans have different risk factors for developing dementia than WA Veterans. Using logistic regression to investigate risk factors for incident dementia in all VHA, we identified different risk factors in older BA and WA Veterans (Cheng et al., 2020). For example, among the key baseline characteristics that were significant predictors of dementia in both races, stroke was a significantly stronger predictor among BAs, and Hispanic ethnicity and depression were significantly stronger predictors among WAs (p<0.0001).Those findings motivated the development of the race-specific risk models proposed in the current study, which instead focuses on prediction.

Many studies have applied NLP and ML methods in dementia (Chang et al., 2021), particularly in the context of neuroimaging (Popuri et al., 2020, Qiu et al., 2020) or in the use of EHRs to identify cognitive impairment or diagnosed dementia (Amra et al., 2017, Wray et al., 2014), yet few studies have sought to use EHRs as a direct phenotyping tool for undiagnosed dementia. Researchers in the UK developed models (including SVM) to identify patients with dementia (Jammeh et al., 2018), and Kaiser Permanente/UCSF researchers developed the eRADAR tool in research participants and then validated it in two health-care systems (Barnes et al., 2020, Coley et al., 2022); both studies limited their EHR interrogations to structured data and have shown some success in identifying undiagnosed dementia. Likewise, Yadgir et al. used ML to identify structured variables associated with cognitive impairment in ER patients (Yadgir et al., 2022). Conversely, Boustani et al. have developed passive digital signatures for ADRD by searching for predetermined variables in *both* structured and unstructured EHR data, and their work suggests that the combination can improve AUC by up to .11 (Boustani et al., 2020); however, like Barnes et al., Boustani et al. use curated, preselected search terms rather than leveraging the potential of supervised ML to identify topic features associated with dementia.

Rather than employing a targeted-word study design like Barnes et al. or Boustani et al., we have sought to improve the identification of dementia by combining supervised ML with an improved clinical standard. More specifically, we have to sought to improve upon EHR ICD codes as the basis for ML by incorporating chart reviews by reviewers who have been blinded to the initial ML-derived dementia likelihood scores. We previously published a ML logistic regression model that used this approach on a smaller scale, applying supervised ML to structured and unstructured data from EHRs to identify topics associated with dementia and then identify cases with undiagnosed dementia (Shao et al., 2019). That study included blinded manual reviews in a smaller sample (n=140) than our current work and produced a sensitivity of 0.825 and a specificity of 0.832. It also had older Veterans (i.e., an average age of 80 vs. 71 in this study); complications with controls in the logistic regression model; and an ad-hoc stratification method for computing sensitivity and specificity, whereas our SVM models avoid these idiosyncrasies in a much larger (n=1,200) and more diverse (600 BA and 600 WA) validation effort.

EHR tools and ML models that do not specifically attempt to reflect minoritized communities are more likely to unintentionally generate cycles of exclusion and to thereby perpetuate underdiagnosis in BAs rather than addressing underdiagnosis (Bracic et al., 2022). To our knowledge the present study is the first effort to develop and evaluate a model that specifically focuses on BAs.

### 4.3 Implications

We seek to develop EHR-based dementia risk scores to support future screening of dementia in clinical settings that include both WAs and BAs. Other researchers have noted that PPV and NPV are better at assessing a screening test in clinical practice than sensitivity and specificity (Trevethan, 2017). Our model generated a very high NPV at the 90^th^ percentile for both BA Veterans (0.98 [0.96, 0.99]) and WA Veterans (0.98 [0.94, 0.99]). These findings are similar to the NPVs reported with the eRADAR tool in an EHR sample that was 89% WA (Barnes et al., 2020) but higher than the NPV reported by Yadgir et al (i.e., 0.93) (Yadgir et al., 2022). The PPV in our study was low for both BA Veterans (0.26 [0.21,0.31]) and WA Veterans (0.15 [0.12,0.20]) at the 90^th^ percentile cutoff. Practically, this means that at that threshold, about a quarter of the BAs and a seventh of the WAs who were flagged by our model as having potential dementia would actually have dementia according to our manual chart reviews. In contrast, Yadgir et al., achieved PPVs greater than 0.4, but to do so, they applied threshold cutoffs higher than 0.8; this meant that they obtained a high true positive rate at the expense of low sensitivity, which is not optimal as a screening instrument given the high cutoff scores (Yadgir et al., 2022). Our algorithms compare very favorably to the eRADAR tool for dementia, which had a PPV of 0.115 in a research setting and 0.020 to 0.048 in patients (Barnes et al., 2020, Coley et al., 2022). Our PPV is similar or superior to the rates of standard screening methods for cancers like mammograms or colonoscopy (reviewed in Barnes et al., 2020). However, cancer screening is often followed by more definitive tests, such as ultrasound and/or biopsies, and thus low PPVs in screening tests may be acceptable. Development of multitier screening and diagnostic tests are therefore necessary prior to the implementation of our SVM model in clinical workflows.

### 4.4 Limitations and future work

The VA patient population skews heavily toward older males, and our training and test data thus had a low percentage of females; that may limit the generalizability of our final ML models outside VHA, though we expect that the same steps could be applied to generate risk scores within other health care systems with more females. In evaluating the low PPVs in our study, it may be that our standards for the diagnosis of dementia (i.e., manual chart review) are flawed, as and that due to insufficient information in the charts, we were unable to retrospectively apply the newest AD criteria (NIA-Reagan) are flawed.{Hyman, 1997, 9329452} If signs and symptoms relevant to impairment are not mentioned in clinical notes, reviewers are unable to assign an dementia diagnosis due to insufficient information. Here, this may have led to a low level of dementia prevalence, and a low prevalence of any condition leads to models with high NPVs and low PPVs. It is possible, therefore, that our model may catch signs of dementia that cannot be captured by a manual chart review, which means our model may perform better when compared to more accurate diagnostic standards, like in-person expert diagnoses or neuropathological assessments; this represents a promising area for future research.

We recognize that future studies need to assess the portability of the ML models that we have developed. Not all EHRs have notes available to researchers (due to privacy issues), and in those instances, researchers will be unable to leverage the full benefit of our models’ ability to search both structured and unstructured data. Future studies should investigate how other ML methods, like deep learning approaches, might improve the detection of undiagnosed dementia; solicit input from BA stakeholders regarding model implementation in clinical processes; and investigate the implementation of our EHR-based risk scores in clinics that serve both BA and WA Veterans.

## Data Availability

The VA EHR data resides in VINCI behind VA firewalls. VA-approved investigators can access the data. SVM algorithms can be made available to interested qualified investigators.

## Acknowledgments

This work was supported by NIA R56 AG059739 and was supported in part by the U. S. Department of Veterans Affairs Office of Research and Development Biomedical Laboratory Research Program.

## Contribution to the field

Up to 50% of dementia is underdiagnosed in primary care settings. This underdiagnosis is especially problematic in elderly Black Americans. Screening all elderly patients (or all elderly Black patients) is time- and resource-intensive, and broad-based screening is not aligned with current clinical guidelines. Thus, other approaches are necessary. This illustrates that cost-effective machine learning algorithms can identify a subset of patients within existing electronic health records who are at high risks for developing dementia. Furthermore, we are one of the first to develop a race-specific algorithm in the context of dementia identification and to thereby leverage machine learning to specifically address dementia-related health-care disparities in Black Americans. That is critical, as models that are not designed to reflect minority population may instead perpetuate underdiagnosis. Moreover, this work may also have tangible financial benefits. Incorporating electronic health records–based algorithms into screening workflows with diagnostic tests as follow-up could focus resources where they will have the most impact in primary care settings, including the prevention of costly health care events that otherwise tend to precede diagnosis.

## Author contributions

Study design (DT, QZ, SM); model development (YS, QZ, DT); data analyses (SM, KT, KB, KW); clinical expertise (DT, AT, KC); initial drafting of the paper and literature review (SM, ASD)

