## Supplemental Table 1 for "Identifying probable dementia in undiagnosed Black and White Americans using machine learning in Veterans Health Administration electronic health records"

**Supplemental Table 1a.** Top 20 most important variables in the support vector machine (SVM) model for Black American (BA) Veterans, ranked by absolute value of variable weight in the model.

| ***Type*** | ***Variable Definition*** | ***Weight*** |
| --- | --- | --- |
| Topic | memory, cognitive, dementia, impairment, wife, loss, problems, donepezil, testing, mild, ... | 0.454 |
| Demographic | Age (normalized to be between 0 and 1 with 0=65yrs and 1=85yrs) | 0.237 |
| Diagnosis | Memory loss/anemia (ICD9: 780.93; ICD10: R41.[1-3]) | 0.144 |
| Topic | weight, move, program, management, obesity, health, loss, bmi, risks, discussed, ... | **- 0.116** |
| Procedure | CT HEAD/BRAIN, W/O DYE (CPT: 70450) | 0.103 |
| Topic | wife, home, reports, states, accompanied, spoke, husband, concerned, house, giving, ... | 0.102 |
| Topic | time, correct, point, work, back, score, points, state, clock, year, ... | 0.092 |
| Topic | point, assistance, points, independently, score, total, shopping, batching, activities, ability, ... | 0.090 |
| Topic | mri, brain, ct, head, acute, small, left, matter, intracranial, white, ... | 0.082 |
| Topic | daughter, mh, home, father, spoke, family, br, states, lives, stated, ... | 0.082 |
| Topic | nursing, rehab, home, placement, facility, care, va, nh, cnh, contract, ... | 0.071 |
| Diagnosis | Mild cognitive impairment (ICD9: 331.83; ICD10: G31.84) | 0.070 |
| Procedure | VITAMIN B-12 (CPT: 82607) | 0.066 |
| Diagnosis | Psychotic disorder with hallucinations (ICD9: 293.82; ICD10: 294.[8,9], F06.[0,8]) | 0.063 |
| Topic | cva, stroke, weakness, sided, left, cerebralvascular, hemiparesis, residual, speech, accident, ... | 0.062 |
| Topic | wife, mrs, spouse, spoke, called, call, husband, stated, mr, home, ... | 0.062 |
| Topic | walker, cane, gait, walking, ambulation, walk, rollator, ambulates, falls, difficulty, ... | 0.058 |
| Topic | tremor, parkinson, tremors, disease, hand, gait, mild, sinemet, pd, levodopa | 0.057 |
| Topic | medication, pill, box, medications, taking, med, pills, meds, bottle, bottles, ... | 0.057 |
| Procedure | SYPHILIS TEST NON-PREP QUAL (CPT: 86592) | 0.056 |

**Supplemental Table 1b.** Top 20 most important variables in the support vector machine (SVM) model for White American (WA) Veterans, ranked by absolute value of variable weight in the model.

| ***Type*** | ***Variable Definition*** | ***Weight*** |
| --- | --- | --- |
| Topic | memory, cognitive, dementia, impairment, wife, loss, problems, donepezil, testing, mild, ... | 0.468 |
| Demographic | Age (normalized to be between 0 and 1 with 0=65yrs and 1=85yrs) | 0.225 |
| Diagnosis | Memory loss/anemia (ICD9: 780.93; ICD10: R41.[1-3]) | 0.180 |
| Topic | wife, home, reports, states, accompanied, spoke, husband, concerned, house, giving, ... | 0.126 |
| Topic | weight, move, program, management, obesity, health, loss, bmi, risks, discussed, ... | **- 0.120** |
| Topic | point, assistance, points, independently, score, total, shopping, batching, activities, ability, ... | 0.117 |
| Topic | mri, brain, ct, head, acute, small, left, matter, intracranial, white, ... | 0.102 |
| Topic | time, correct, point, work, back, score, points, state, clock, year, ... | 0.100 |
| Topic | tremor, parkinson, tremors, disease, hand, gait, mild, sinemet, pd, levodopa, ... | 0.087 |
| Procedure | CT HEAD/BRAIN, W/O DYE (CPT: 70450) | 0.082 |
| Topic | daughter, mh, home, father, spoke, family, br, states, lives, stated, ... | 0.079 |
| Diagnosis | Psychotic disorder with hallucinations (ICD9: 293.82, 294.[8,9]; ICD10: F06.[0,8]) | 0.072 |
| Topic | nursing, rehab, home, placement, facility, care, va, nh, cnh, contract, ... | 0.069 |
| Topic | walker, cane, gait, walking, ambulation, walk, rollator, ambulates, falls, difficulty, ... | 0.067 |
| Diagnosis | Mild cognitive impairment (ICD9: 331.83; ICD10: G31.84) | 0.065 |
| Topic | memory, average, cognitive, test, range, impaired, functioning, performance, evaluation, testing, ... | 0.064 |
| Diagnosis | Parkinson's disease (ICD9: 332.0; ICD10: G20., G21.4) | 0.062 |
| Topic | injury, head, fall, fell, hit, back, ago, fracture, rib, trauma, ... | 0.058 |
| Procedure | SYPHILIS TEST NON-PREP QUAL (CPT: 86592) | 0.057 |
| Procedure | VITAMIN B-12 (CPT: 82607) | 0.055 |
