## Supplemental Table 2 for "Identifying probable dementia in undiagnosed Black and White Americans using machine learning in Veterans Health Administration electronic health records"

**Supplemental Table 2.** Values of sensitivity, specificity, positive predictive value (PPV), and negative predictive value (NPV) at various score cutoffs. Values in brackets denote 95% confidence intervals.^*^

| **Race**^†^ | **Cutoff Percentile** | **Sensitivity** | **Specificity** | **PPV** | **NPV** |
| --- | --- | --- | --- | --- | --- |
| BA | 50 | 0.89 [0.53, 1] | 0.48 [0.42, 0.54] | 0.08 [0.06, 0.10] | 0.99 [0.93, 1] |
|  | 75 | 0.89 [0.53, 1] | 0.76 [0.76, 0.77] | 0.15 [0.12, 0.19] | 0.99 [0.95, 1] |
|  | 90 | 0.61 [0.40, 0.76] | 0.92 [0.91, 0.92] | 0.26 [0.21, 0.31] | 0.98 [0.96, 0.99] |
|  | 95 | 0.37 [0.24, 0.48] | 0.97 [0.96, 0.97] | 0.31 [0.25, 0.39] | 0.97 [0.95, 0.98] |
| WA | 50 | 0.86 [0.47, 1] | 0.51 [0.45, 0.57] | 0.06 [0.04, 0.11] | 0.99 [0.94, 1] |
|  | 75 | 0.58 [0.31, 0.85] | 0.76 [0.76, 0.77] | 0.09 [0.06, 0.12] | 0.98 [0.93, 0.99] |
|  | 90 | 0.43 [0.24, 0.67] | 0.91 [0.91, 0.92] | 0.15 [0.12, 0.20] | 0.98 [0.94, 0.99] |
|  | 95 | 0.30 [0.16, 0.48] | 0.96 [0.96, 0.96] | 0.22 [0.16, 0.29] | 0.97 [0.94, 0.99] |

^*^ Estimates were computed using inverse probability weighting to account for stratified sampling, and confidence intervals were computed using bootstrapping.

^†^ BA: Black American; WA: White American.
