## Supplemental Figure 1 for "Identifying probable dementia in undiagnosed Black and White Americans using machine learning in Veterans Health Administration electronic health records"

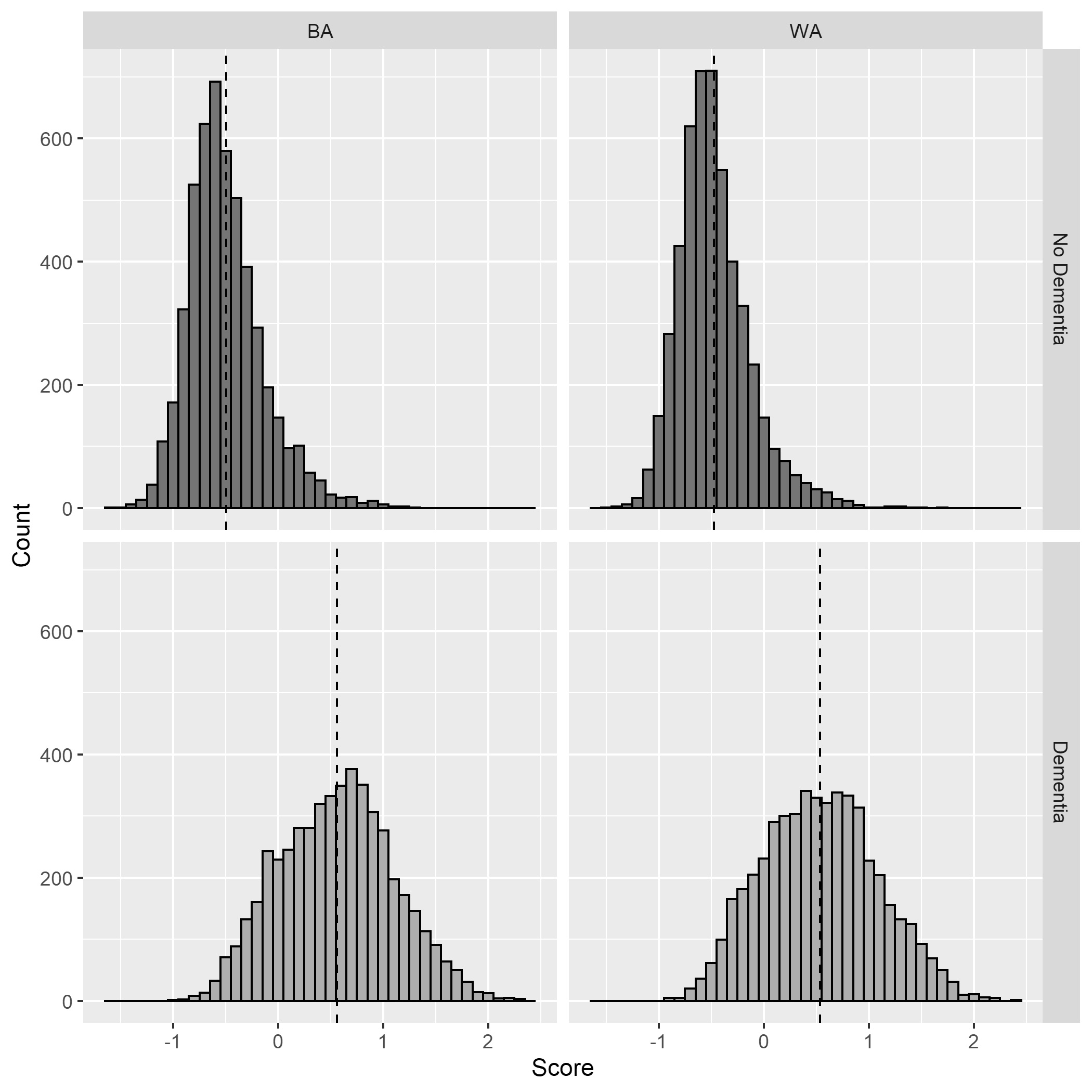

**Supplemental Figure 1.** Distribution of scores by dementia status and race (BA: Black American; WA: White American) for Veterans in the Training Sample (n = 5,000 in each dementia status group for each race). Dashed lines represent the means of the distribution.
