## Supplemental Figure 2 for "Identifying probable dementia in undiagnosed Black and White Americans using machine learning in Veterans Health Administration electronic health records"

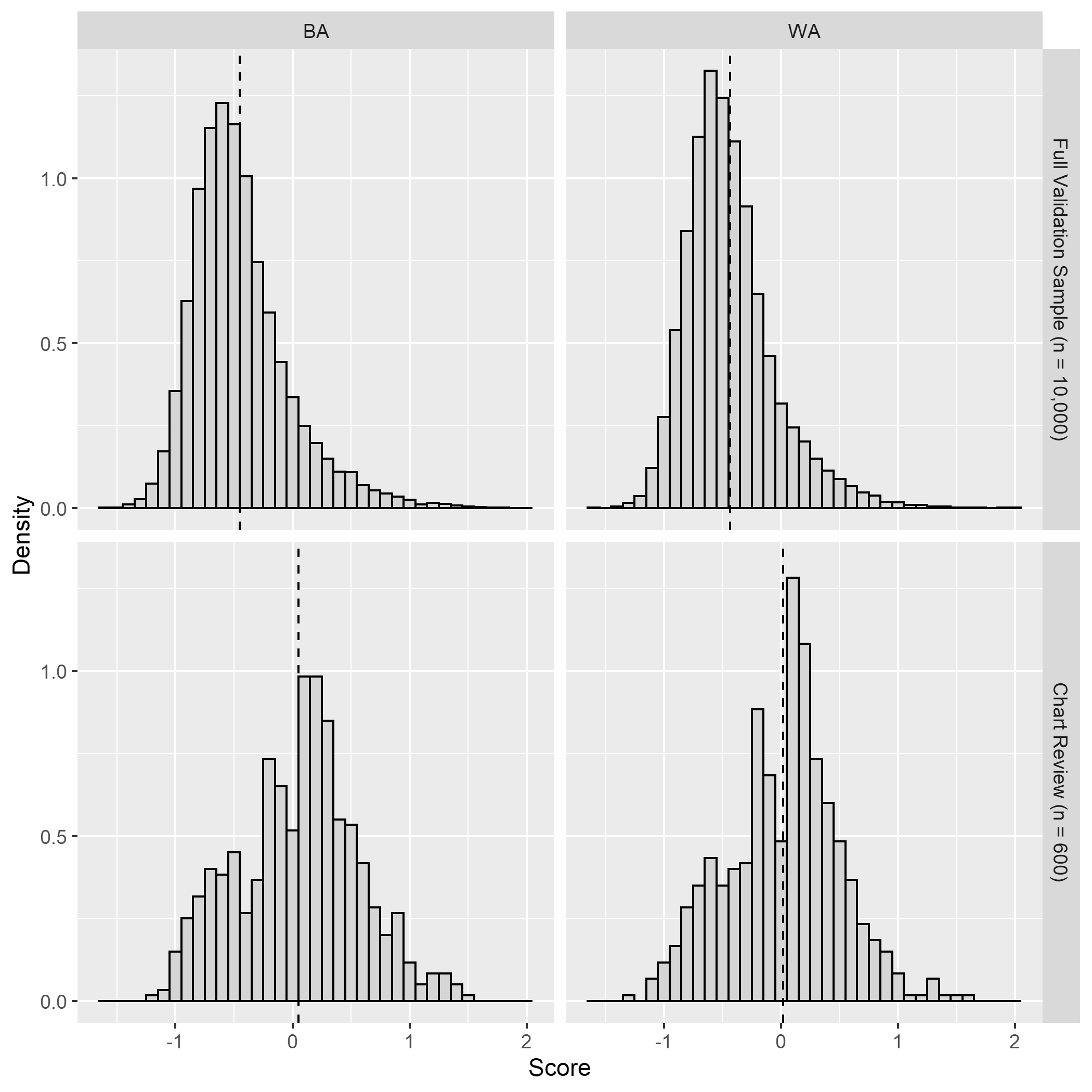

**Supplemental Figure 2.** Distribution of scores by race (BA: Black American; WA: White American) for Veterans in the Validation Sample (n = 10,000 for each race; n = 600 had chart review for each race). Dashed lines represent the distribution means.
