## Supplemental Figure 3 for "Identifying probable dementia in undiagnosed Black and White Americans using machine learning in Veterans Health Administration electronic health records"

**BA**

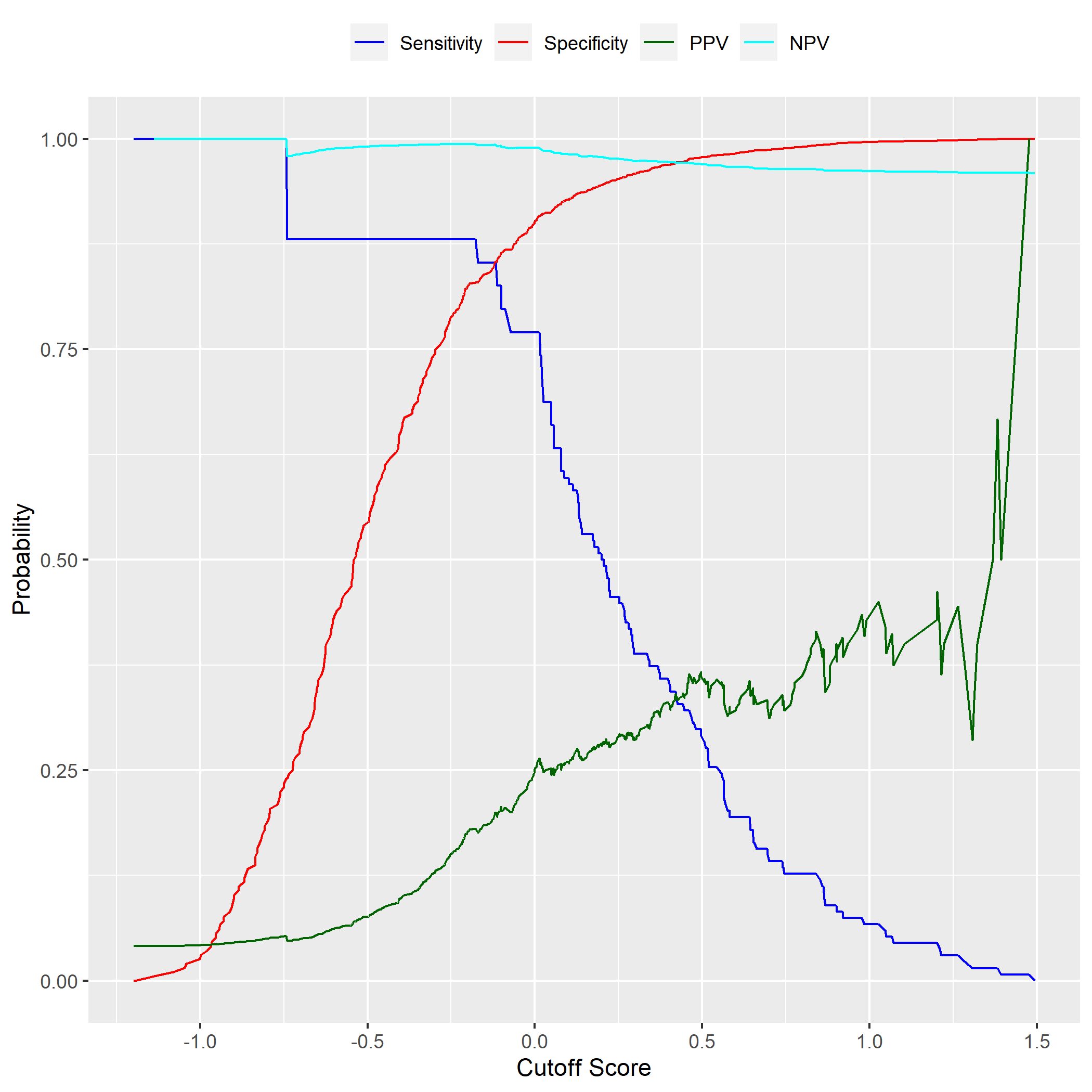

**WA**

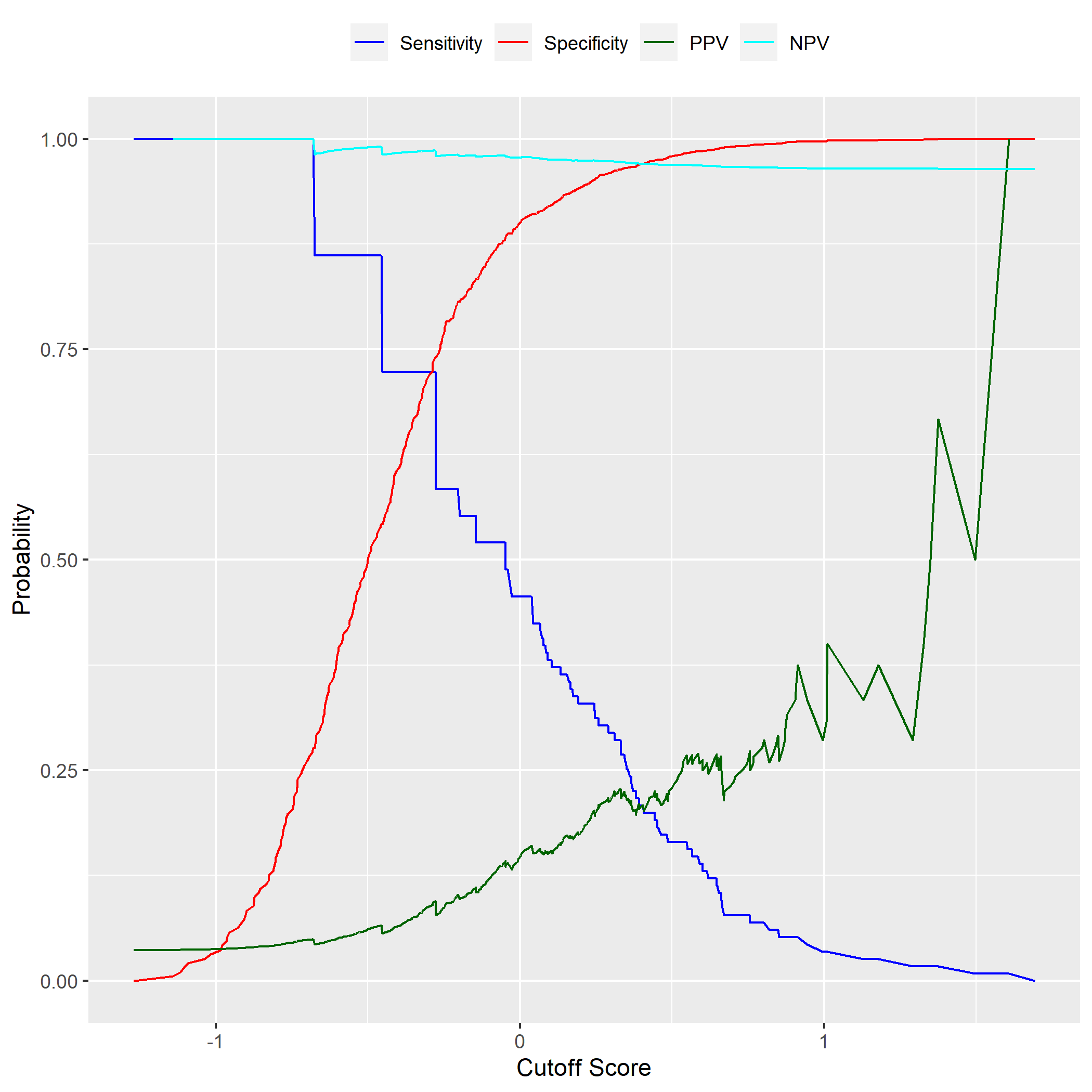

**Supplemental Figure 3.** Observed values of sensitivity, specificity, positive predictive value (PPV), and negative predictive value (NPV) versus score cutoff by race (BA: Black American; WA: White American) based on the Veterans who had chart review.
