## Supplemental Figure 4 for "Identifying probable dementia in undiagnosed Black and White Americans using machine learning in Veterans Health Administration electronic health records"

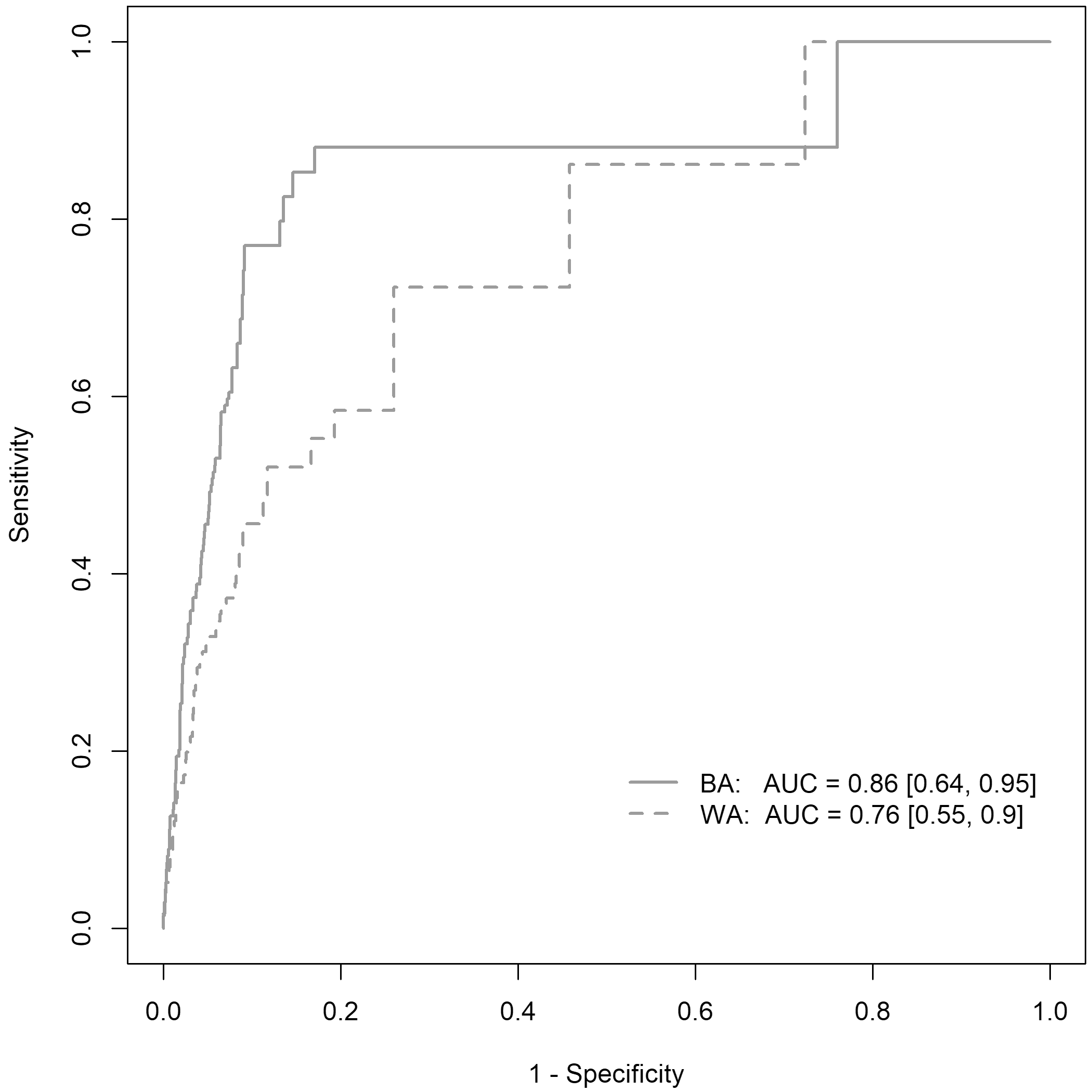

**Supplemental Figure 4.** Receiver operating characteristic (ROC) curves based on observed values of sensitivity and specificity for the Black American (BA) and White American (WA) Veterans who had chart reviews. Values in brackets represent the 95% confidence interval for the area under the curve (AUC).
